# A systematic review and meta-analysis of randomised controlled trials examining the effect of ultra-processed food on energy intake and weight gain

**DOI:** 10.64898/2026.06.03.26354787

**Authors:** Eric Robinson, Andrew Jones, Rebecca Evans, Amy Finlay, Jane Brealey, Thomas Gough, Jenna Cummings, Esther Fisher, Madeleine Jutla, Euba Morenikeji Ibilola, Victoria Norton

## Abstract

Ultra-processed food (UPF) may contribute to increased energy intake and weight gain, but evidence synthesis from randomised controlled trials (RCT) is lacking. A pre-registered systematic review and meta-analysis of RCTs was conducted comparing UPF with less processed food (LPF) on energy intake and/or body weight in humans. Secondary analyses (meta-regression and sub-group) examined effects of UPF on appetite sensations, eating rate, palatability and considered the role of nutrient profile in explaining results. Ten eligible studies were included. UPF trial arms tended to have higher energy intake (standardised mean differences [SMDs]=0.18-0.44), but statistical significance varied between analytic models. Weight gain (SMD=0.65) and eating rate (SMD=0.96) were significantly greater in UPF trial arms. No significant differences in palatability, appetite sensations or energy intake later in the day were observed. Diets (UPF vs. LPF) used in trials were not matched for nutrient profile. Effects on energy intake varied if UPFs were higher (SMD=0.71) or similar (SMD=0.02) in energy density. Current RCTs are suggestive that UPFs may increase energy intake and body weight; however, results may be explained by energy density of foods used. Further research is needed to understand whether the level of processing impacts health outcomes independent to nutrient profile.

## 1. Introduction

Ultra-processed foods (UPFs) have been characterised as food products which are formulations of ingredients, mostly of exclusive industrial use, that result from a series of industrial processes (Monteiro et al., 2019). Based on the Nova classification, UPFs make a considerable contribution to dietary intakes in many countries (Marino et al., 2021). Observational research has linked higher consumption of UPFs to a broad range of negative health outcomes (Lane et al., 2024). In particular, UPF consumption has been linked to higher energy intake (Lane et al., 2024) and weight gain (Crimarco et al., 2022). These outcomes are particularly important because obesity is a risk factor for further non-communicable disease (Nyberg et al., 2018) and weight gain associated with increased UPF consumption may in part explain other UPF-health outcome relationships, such as development of cancer (Kong et al., 2025; Morales-Berstein et al., 2024).

There is scientific uncertainty over why diets higher in UPF are associated with health outcomes (Robinson & Johnstone, 2024) and due to a range of limitations inherent to observational research (e.g., such as confounding (Robinson & Jones, 2024)), randomised controlled trials (RCTs) are required to provide causal evidence (O’Connor et al., 2023). Until recently there was only one RCT (Hall et al., 2019) examining the effect of a diet high in UPF with a less processed food (LPF) and both daily energy intake and 2-week weight gain were higher during the UPF diet (Hall et al., 2019). More recent RCTs in which the effects of UPF vs. LPF on energy intake and/or body weight were compared have produced more mixed findings (Dicken et al., 2025; Larcom et al., 2026; Lasschuijt et al., 2023).

A range of potential explanations for why diets higher in UPF may promote weight gain have been proposed, including UPFs displacing other foods from the diet or presence of additives and/or flavourings or disruption of food matrices that increase palatability and eating rate and decrease satiating effects (Juul et al., 2025; Monteiro et al., 2025). Another possible explanation for why UPF consumption is associated with negative health outcomes is the nutrient profile of many UPFs (Mendoza & Hu, 2025). Compared to LPF, diets rich in UPFs tend to be higher in saturated fat, salt and/or sugar, and lower in fibre (Martini et al., 2021). These differences in nutrient profile result in many UPFs having a higher energy density than some LPF (Gupta et al., 2019). Energy density is a known driver of daily energy intake and body weight change (Robinson et al., 2022; Rolls, 2009). In the Hall et al. (2019) trial the non-beverage UPFs provided to participants were higher in energy density than the non-beverage LPF (2.15 vs 1.15 kcal/g); therefore, confounded with degree of processing (Fazzino et al., 2023). To date, evidence synthesis from RCTs is lacking on whether ultra processing itself acts as a driver of overeating independent to nutritional profile. Answering this question will have direct implications for public health measures to improve diet as well as provide valuable insights for future RCTs (Cullum, 2024; Robinson & Johnstone, 2024).

This present research conducted a systematic review and meta-analysis of RCTs on UPF compared with LPF on study outcomes. The first objective pooled all experimental studies quantifying effects of manipulating UPF consumption on energy intake and/or body weight (primary outcomes), alongside relevant secondary outcomes (e.g., appetite sensations, eating rate, palatability, etc.). The second objective examined whether there is evidence from RCTs that UPF consumption affects energy intake and/or weight gain independent to, or is more likely explained by, nutrient profile, particularly energy density. Given the use of RCT designs to understand health effects of UPF is an emerging science, we also aimed to characterise other types of health outcomes examined in RCTs and provide recommendations for future studies.

## 2. Methods

The review was pre-registered on PROSPERO (CRD420251183053) and Open Science Framework (https://osf.io/sdrcz) and conducted according to PRISMA guidelines (Page et al., 2021).

### Eligibility Criteria

Participants: Studies of human participants, with no exclusions based on participant characteristics (e.g., age).

Intervention: Studies in which participants consumed an increased amount of UPF (relative to a comparison condition) during a meal or across multiple days, achieved through either researcher provision of food and/or dietary guidance. UPF defined using Nova classification or an alternative validated classification system.

Comparison: Studies in which participants consumed a decreased amount of UPF (relative to intervention condition) during a meal or across multiple days.

Primary Outcomes: Studies measuring ad-libitum energy intake (e.g., meal or daily intake) and/or body weight change.

Context: Studies with parallel arms (between-subjects) and/or cross-over (within-subjects) study designs, including laboratory experiments. Because review objectives concern the effects of increased vs. decreased UPF consumption on outcomes, studies not providing evidence of differences in UPF consumption between trial arms (e.g., no significant difference in daily intake reported) were ineligible.

### Information Sources

Search strategy: Electronic database searches of PUBMED, SCOPUS and Cochrane Central Register were conducted November 2025 with no language restrictions (primary search strategy). We searched from 2009 onwards as this was when the Nova classification system was introduced. We searched combinations of terms relating to UPFs/food processing and primary outcomes of interest. See supplementary materials for full search terms. To identify further potentially eligible articles, Google Scholar, medRxiv and the Open Science Framework pre-print servers were searched (published journal articles and published pre-prints were both eligible). During the search process, if we identified trial protocol papers for studies that may have been eligible, we emailed the trial’s point of contact to inquire about availability of study results in a published journal or pre-print article. Reference lists of eligible studies were searched.

### Selection Process and Extraction

For title and abstract screening of primary database searches, a minimum of two authors independently assessed each record, and discrepancies were resolved through discussion. A single author conducted supplementary searches to identify potentially eligible articles eligible for full-text screening. Two researchers independently completed full-text screening with disagreements resolved through discussion and/or a third researcher (rationale for study exclusion outlined in supplementary materials).

A minimum of two researchers independently extracted information from eligible articles. Any minor inconsistencies were resolved through discussion. Authors of eligible articles were contacted for missing statistical data required for meta-analysis of primary outcomes and where possible, we also used WebPlotDigitizer (version 5.2) to extract missing secondary outcome data from eligible articles. We extracted information on population sampled, study setting, design, procedures and duration. Nature of intervention vs. control group delivery (e.g., food provision vs. dietary guidance) and their characterisation (e.g., amount of UPF vs. other food sources, macronutrient profile and energy density of foods) were extracted alongside primary outcome data (energy intake, body weight) and results. We favoured extraction of energy intake outcome data for longest duration possible (e.g., average daily energy intake over full trial duration) and extraction of weight change (kg) by trial end. We extracted data on a limited number of secondary outcomes which could differ between trial arms and explain why UPF increases energy intake and/or body weight: food palatability, eating rate, appetite sensation. A small number of studies compared effects of intervention vs. control diets on subsequent energy intake (e.g., ad-libitum energy intake for the rest of the day after consuming UPFs), so we extracted individual study results relevant to this outcome. We recorded other types of health-relevant outcomes collected in studies (e.g., metabolic biomarkers) for descriptive purposes.

### Risk of Bias

For objective one (effect of UPF on outcomes), two researchers independently assessed risk of bias through the use of NUQUEST – NUtrition QUality Evaluation Strengthening Tool for randomized control trials (Kelly et al., 2022). Disagreements were resolved through discussion. For the NUQUEST item ‘*the only difference between groups is the intervention under investigation*’ we did not consider differences in nutritional profile as indicative of high risk of bias when broadly considering the effect of UPF on outcomes (objective one). For objective two (evidence for effect of processing independent to nutritional profile), we did consider meaningful differences (i.e., 10% difference tolerance) between trial arms for key nutrients (i.e., carbohydrates, sugar, fats, protein, salt, fibre, energy density) as being indicative of high risk of bias. If a study did not report the above information or reported data consistent with meaningful differences (>10%) between intervention and control diets, it was considered unsuitable to draw conclusions from for objective two.

### Data Synthesis

#### Pooled effect of UPF on energy intake and weight gain

We conducted meta-analysis for primary and secondary outcomes in which there was a minimum of three eligible studies using the ‘metafor’ package in R (version 4.5.2). Because there was variability in duration (e.g. weight change over days vs. weeks) and measurement (e.g., single meal energy intake vs. daily energy intake) we conducted primary analyses with a standard measure of effect size (standardised mean difference: SMD) and raw outcomes (kg, kcals), for descriptive purposes. Studies could contribute multiple effect sizes to meta-analyses, so a random intercept for study ID was included, where appropriate. For full information on effect size computation, see supplementary materials. We adopted a range of approaches to examine potential publication bias and the extent to which results were dependent on individual studies (see supplementary materials for full details).We used the same analytic approach to examine secondary outcomes.

#### Effect of processing independent to nutritional profile

In studies providing participants with food and examining effects on energy intake, differences between intervention vs. control diets for individual nutrients were variable, resulting in variability in magnitude of intervention vs. control diet arm energy density differences. We conducted formal sub-group analyses comparing trial arms in which the energy density of UPF diets were markedly higher than corresponding LPF (diets (>10%) vs. trial arms in which energy density of UPFs was similar or lower (<10%) than LPF. We selected a 10% threshold for markedly higher a-priori, as we assumed closer matching (<5%) would be unlikely to achieve in trials. We also meta-regressed size of energy density difference between UPF vs. LPF diets in each study with energy intake difference between trial arms to examine if variations in energy density of diets used explained results.

### Reporting Bias and Certainty of Evidence

We used a range of methods and sensitivity analyses to assess evidence of potential publication bias and influential cases, for full details see supplementary materials. Grading of Recommendations Assessment, Development and Evaluation (GRADE) was used to classify evidential certainty as high, moderate, low and very low.

## 3. Results

### Studies included

Ten studies were included (de Oliveira et al., 2025; Dicken et al., 2025; Discepoli et al., 2026; Hall et al., 2019; Hamano et al., 2024; Larcom et al., 2026; Lasschuijt et al., 2023; Preston et al., 2025; Rego et al., 2026; Teo et al., 2022). See Figure 1 for search process results and supplementary materials for common exclusions. Three studies were conducted in the US (Hall et al., 2019; Larcom et al., 2026; Rego et al., 2026), four in Europe (Dicken et al., 2025; Discepoli et al., 2026; Lasschuijt et al., 2023; Preston et al., 2025) and individual studies were conducted in Brazil (de Oliveira et al., 2025), Singapore (Teo et al., 2022) and Japan (Hamano et al., 2024). Three studies sampled participants with overweight or obesity (de Oliveira et al., 2025; Dicken et al., 2025; Hamano et al., 2024). Studies adopted cross-over (n = 7; Hall et al., 2019; Hamano et al., 2024; Larcom et al., 2026; Lasschuijt et al., 2023; Teo et al., 2022; Dicken et al., 2025; Rego et al., 2026), parallel arms (n = 2; de Oliveira et al., 2025; Discepoli et al., 2026) and mixed (n = 1; Preston et al., 2025) designs. Five studies were conducted in laboratory settings (Hall et al., 2019; Hamano et al., 2024; Larcom et al., 2026; Lasschuijt et al., 2023; Teo et al., 2022) and five in real-world settings (de Oliveira et al., 2025; Dicken et al., 2025; Discepoli et al., 2026; Preston et al., 2025; Rego et al., 2026). The majority of studies provided participants with UPF vs. LPF meals to consume (8 out of 10) (Dicken et al., 2025; Hall et al., 2019; Hamano et al., 2024; Larcom et al., 2026; Lasschuijt et al., 2023; Preston et al., 2025; Rego et al., 2026; Teo et al., 2022) and two studies (de Oliveira et al., 2025; Discepoli et al., 2026) provided participants with dietary advice to reduce UPF consumption. Dietary provision studies typically provided participants with meals which only contained UPF or were predominantly UPF (e.g., 81% energy from UPF) compared to meals consisting of only minimally processed food or predominantly minimally processed food (e.g., 2% energy from UPF), although this was not consistently reported. See tables 1-4 for detailed individual study information.

**Figure 1.**
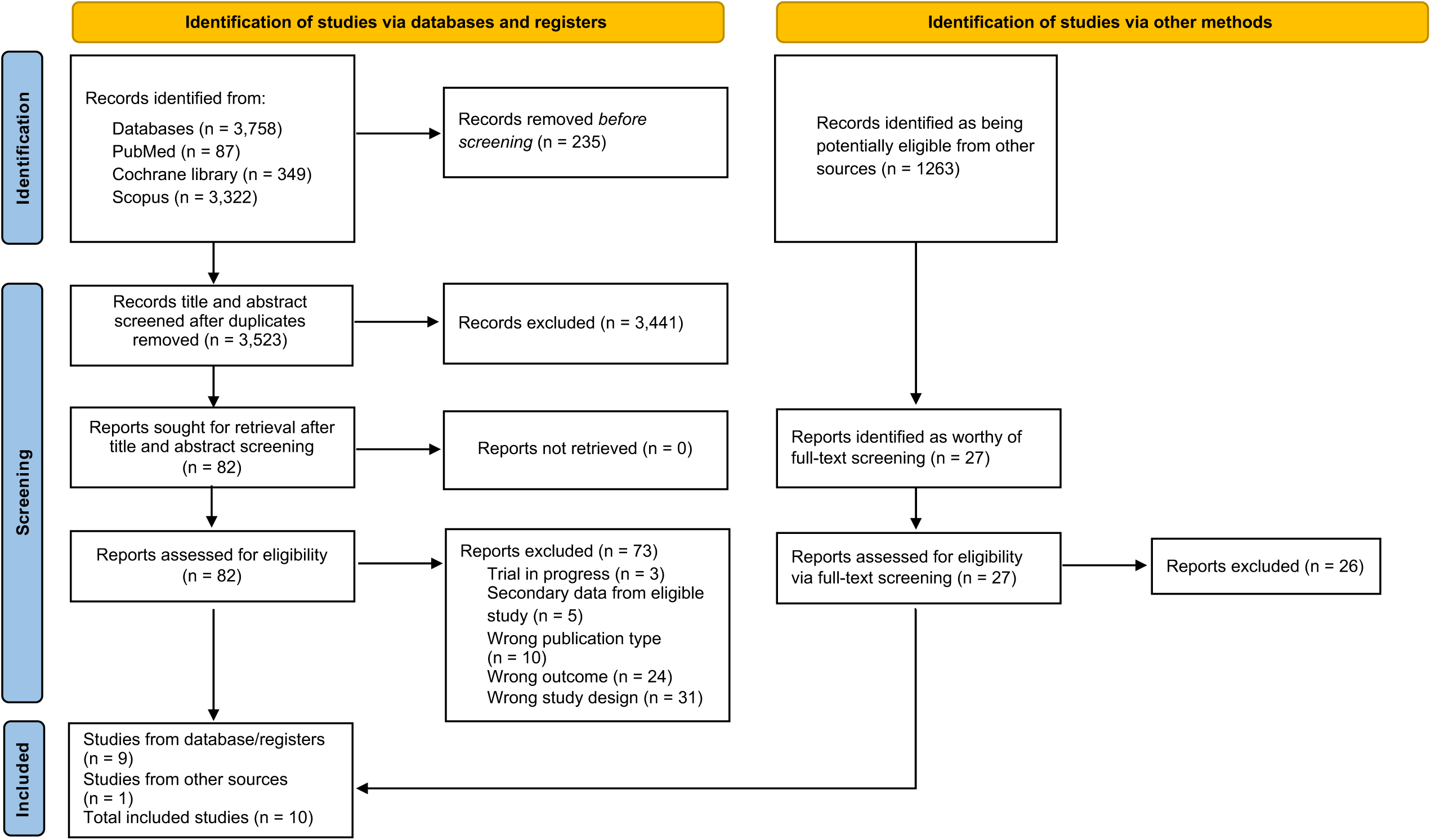
Article identification and inclusion flow chart.

**Table 1.** Design and primary outcome information for included laboratory studies.

| Study | Sample | Study design | Intervention arm(s) | Control arm | Reporting of nutrient composition matching | Primary outcomes and length of assessment | Primary outcome results |
| --- | --- | --- | --- | --- | --- | --- | --- |
| Hall et al. (2019) | 20 US adults<br>10M, 10F<br>M age = 31<br>M BMI = 27 | Cross-over laboratory study<br>All food provided to participants for 2 weeks | Provision of diet higher in UPF<br>81% of energy from UPF for daily meals and snacks<br>N =20 | Provision of diet lower in UPF (0% of energy from UPF for daily meals and snacks)<br>Nova category 1 primary source of energy (88%)<br>N = 20 | Beverage and food described as matched<br>Food described as not matched | Average daily energy intake, researcher measured (2 weeks)<br>Body weight change, researcher measured (2 weeks) | Significant difference between arms for both outcomes<br>**Energy intake (kcal; M ± SD)**<br>Intervention: 2978 ± 997<br>Control: 2470 ± 789<br>**Weight change (kg; M ± SD)**<br>Intervention: 0.93 ± 1.44<br>Control: -0.91 ± 1.34 |
| Hamano et al. (2024) | 9 Japanese adults (overweight/obese)<br>9M, 0F<br>M age = 29<br>M BMI = 27 | Cross-over laboratory study<br>All food provided to participants for 1 week | Provision of diet higher in UPF<br>99% of energy from UPF for daily meals and snacks<br>N = 9 | Provision of diet lower in UPF (0% of energy from UPF for daily meals and snacks)<br>Nova category primary source of energy not reported<br>N = 9 | Meals described as being matched | Average daily energy intake, researcher measured (1 week)<br>Body weight change, researcher measured (1 week) | Significant difference between arms for both outcomes<br>**Energy intake (kcal; M ± SE)**<br>Intervention: 4126 ± 229<br>Control: 3313 ± 302<br>**Weight change (kg; M ± SE)**<br>Intervention: 2.18 ± 0.43<br>Control: 1.07 ± 0.58 |
| Larcom et al. (2026) | 40 US adults<br>28F, 12M<br>M age = 25<br>M BMI = 25 | Cross-over laboratory study<br>Single meal provided to participants | (A) Provision UPF-based breakfast ‘high nutritional quality’<br>(B) Provision UPF-based breakfast ‘low | Provision of LPF breakfast ‘high nutritional quality’<br>Nova category primary source of energy not reported | ‘High nutritional quality’ UPF vs. non-UPF described as matched<br>‘Low nutritional quality’ UPF and ‘high nutritional quality’ non-UPF described as not matched | Energy intake from single meal, researcher measured<br>Subsequent energy intake (remainder of the day) | Significant energy intake difference between intervention B and control, but not intervention A and control<br>**Energy intake (kcal; M ± SD)**<br>Intervention (A): 610 ± 152<br>Intervention (B): 503 ± 164<br>Control: 586 ± 163 |
|  |  |  | nutritional quality'<br>100% of energy from UPF<br>N=40 | N=40 |  |  | Subsequent energy intake measured during remainder of the day did not differ between intervention vs. control arms (data not reported) |
| Lasschuijt et al. (2023) | 18 Dutch adults<br>7 M, 11F<br>M age = 23<br>M BMI = 22 | Cross-over laboratory study<br>All food provided to participants for 1 day | (A) Provision UPF-based soft meals<br>(B) Provision UPF-based hard meals<br>A=52% and B=29% of energy from UPF<br>N = 18 | (A) Provision LPF-based soft meals<br>(B) Provision LPF-based hard meals<br>Nova category 1 primary source of energy (A=81%, B=84%)<br>N=18 | Meals described as matched | Energy intake across 1 day, researcher measured | No significant energy intake difference between intervention vs. control arms<br>**Energy intake (kcal; M ± SE)**<br>Intervention (A): 2608 ± 136<br>Intervention (B): 1706 ± 139<br>Control (A): 2205 ± 137<br>Control (B): 1999 ± 134 |
| Teo et al. (2022) | 50 Singapore adults<br>24M, 26F<br>M age = 24<br>M BMI = 21 | Cross-over laboratory study<br>Single breakfast meal provided to participants | (A) Provision UPF-based lunch soft in texture<br>(B) Provision UPF-based lunch hard in texture<br>100% of energy from UPF for A & B<br>N = 50 | (A) Provision LPF lunch soft in texture<br>(B) Provision LPF lunch hard in texture<br>Nova category 1 primary source of energy (100%) for A & B<br>N = 50 | Meals described as matched | Energy intake from single meal, researcher measured<br>Subsequent energy intake (remainder of the day) | Significant energy intake difference between intervention (A & B) vs. control arms (A & B)<br>**Energy intake (kcal; M ± SE)**<br>Intervention (A): 789 ± 31.6<br>Intervention (B): 628 ± 26.3<br>Control (A): 724 ± 28.9<br>Control (B): 483 ± 25.5<br>Subsequent energy intake measured during remainder of the day did not differ between intervention vs. control arms (data not reported) |

Abbreviations: M = mean, SD = standard deviation, SE = standard error, N = number of participants, BMI = body mass index, M = males, F = females, UPF = ultra-processed food. LPF = less processed food.

**Table 2.** Design and primary outcome information for real-world studies.

| Study | Sample | Study design | Intervention arm(s) | Control arm | Reporting of nutrient composition/guidance matched | Primary outcomes and length of assessment | Primary outcome results |
| --- | --- | --- | --- | --- | --- | --- | --- |
| Dicken et al. (2025) | 50 UK adults (ITT analysis)<br>5M, 45F<br>M age = 43<br>M BMI = 33 | Cross-over intervention study<br>All food provided to participants for 8 week periods | Provision of diet higher in UPF (91% kcals from UPF)<br>N = 50 | Provision of diet with minimal UPF (2% kcals from UPF)<br>Nova category 1 primary source of energy (82%)<br>N = 50 | Diet provision reported as being matched for adherence to dietary guidelines | Average daily energy intake, self-reported (8 weeks)<br>Body weight change, researcher measured (8 weeks) | Significant difference (increased) between intervention vs. control arm for energy intake and weight change (gain)<br>**Energy intake (kcals; M ± SE)**<br>Intervention: 1745 ± 64.0<br>Control: 1420 ± 92.1<br>**Weight change (kg; M ± SE)**<br>Intervention: -1.59 ± 0.3<br>Control: -2.54 ± 0.4 |
| Discepoli et al. (2026) | 66 Italian young adults<br>32 M, 34F<br>M age = 23<br>M BMI = 23 | Parallel arms intervention study<br>Dietary guidance provided to participants | Dietary advice to reduce consumption of processed food<br>N = 33 | No dietary advice<br>N = 33 | N/A due to non-active control arm | Average daily energy intake, self-reported (16 weeks) | No significant difference between arms<br>**Energy intake (kcals; M ± SD)**<br>Intervention: 2289 ± 672<br>Control: 2124 ± 882 |
| Preston et al. (2025) | 43 Danish adults<br>43M, 0F<br>Age = 20-35<br>BMI =18.5-30 | Cross-over intervention study (mixed design)<br>All food provided to participants for 3 week periods | Provision of diet higher in UPF<br>77% kcals from UPF either:<br>(A) Caloric adequate, N = 21<br>(B) Caloric excessive, N = 22 | Provision of diet with minimal UPF (<1% kcals from UPF)<br>Nova category 1 primary source of energy (66%)<br>(A) Caloric adequate, N = 21<br>(B) Caloric excessive, N = 22 | Diet provision reported as being matched | Body weight change, researcher measured (3 weeks) | Significant difference (greater weight gain) between intervention vs. control arm irrespective of caloric provision (A vs. B)<br>**Weight change (kg; M ± SE)**<br>Intervention (A): 0.17 ± 0.37<br>Intervention (B): 0.33 ± 0.35<br>Control (A): -1.24 ± 0.34<br>Control (B): -0.95 ± 0.34 |
| Rego et al. (2026) | 27 US adolescents and young adults<br>10M, 17F<br>M age = 22<br>M BMI = 24 | Cross-over intervention study | Provision of diet higher in UPF (81% kcals from UPF)<br>N = 27 | Provision of diet with minimal UPF (0% kcals from UPF)<br>Nova category primary source of energy not reported<br>N = 27 | Diet provision reported as being matched for energy and macronutrients | Subsequent ad-libitum meal intake (after 14 day of diets) | No significant difference between arms (M difference: 4.23; d = 0.03) |
| de Oliveira et al. (2025) | 34 Brazilian adults with obesity<br>Gender Not reported<br>M age = 31<br>M BMI = 31 | Parallel arms intervention study<br>Dietary guidance provided to participants | Energy restriction focusing on reducing UPF<br>N = 18 | Energy restriction focusing on reducing habitual food portion sizes and interpreting nutritional information<br>N = 16 | Dietary guidance reported as being matched | Body weight change, researcher measured (6 months) | Significant difference (greater weight gain) between intervention vs. control arm<br>**Weight change (kg; M ± SE)**<br>Intervention: 1.06 ± 0.93<br>Control: -3.41 ± 1.01 |

Abbreviations: M = mean, SD = standard deviation, SE = standard error, N = number of participants, BMI = body mass index, M = males, F = females, UPF = ultra-processed food. LPF = less processed food, ITT = intention-to-treat.

**Table 3.** Additional methodological and secondary outcome information for included laboratory studies.

| Study | Difference in UPF consumption by arm | Intervention arm served food ED (kcal/g) | Control arm served food ED (kcal/g) | Intervention vs control arm food nutrient profile comparisons | Liking assessment | Eating rate assessment | Appetite assessment | Other health outcomes assessed |
| --- | --- | --- | --- | --- | --- | --- | --- | --- |
| Hall et al. (2019) | Not reported<br>Trial design ensures large difference | Food ED = 2.15<br>Food and drink ED = 1.25 | Food ED = 1.15<br>Food and drink ED = 1.13 | Daily meals and snacks:<br>≈Fat<br>>Sat fat<br>≈Carbohydrate<br>>Sugars<br>>Sodium<br><Protein<br><Fibre | Self-reported liking (VAS; 0-100) after first bite of meals<br>No significant difference between arms<br>**Liking (M ± SD)**<br>Intervention: 67.0 ± 10.6<br>Control: 62.8 ± 13.7 | Meal duration recorded by researcher and food intake divided by meal duration (kcal/minute)<br>Significant difference between arms, intervention arm higher eating rate<br>**Eating rate (M ± SD)**<br>Intervention: 49.2 ± 6.9<br>Control: 32.2 ± 4.1 | Hunger and fullness self-reported VAS at multiple time points each day (0-100)<br>No significant difference between arms<br>**Hunger (M ± SD)**<br>Intervention: 35.5 ± 12.4<br>Control: 37.7 ± 14.3<br>**Fullness (M ± SD)**<br>Intervention: 58.1 ± 12.0<br>Control: 56.4 ± 16.5 | Markers of cardiometabolic health<br>Appetite hormones |
| Hamano et al. (2024) | Not reported<br>Trial design ensures large difference | Food ED = 2.0<br>Food and drink ED = 1.7 | Food ED = 1.1<br>Food and drink ED = 1.1 | Daily meals and snacks:<br>≈Fat<br>>Sat fat<br>≈Carbohydrate<br>>Sugars<br>>Sodium<br>≈Protein<br><Fibre | Not measured | Meal duration recorded by researcher and food intake divided by meal duration (kcal/minute)<br>Significant difference between arms, intervention arm higher eating rate<br>**Eating rate (M ± SD)**<br>Intervention: 61.9 ± 18.6<br>Control: 43.0 ± 11.4 | Hunger and fullness self-reported VAS before meals (0-100)<br>No significant differences between arms<br>**Hunger (M ± SD)**<br>Intervention = 54.9 ± 10.2<br>Control = 54.1 ± 12.0<br>**Fullness (M ± SD)**<br>Intervention = 57.8 ± 8.4<br>Control = 56.4 ± 11.9 | Markers of cardiometabolic health<br>Appetite hormones |
| Larcom et al. (2026) | Not reported<br>Trial design ensures large difference | (A) 'High nutritional quality'<br>Food and drink ED = 1.12<br>Food ED = 1.66 | Food and drink ED = 1.13<br>Food ED = 1.64 | (A) vs. Control:<br>≈Fat<br>Sat fat (NR)<br>≈Carbohydrate<br>≈Sugars<br>Sodium (NR)<br>≈Protein<br>>Fibre<br>(B) vs. Control: | Self-reported liking (VAS; 0-100) after meals<br>Intervention B, but not A differed significantly (lower liking) to Control<br>**Liking (M ± SD)**<br>Intervention (A): 65.3 ± 22.9<br>Intervention (B): 45.4 ± 20.9<br>Control: 53.0 ± 24.4 | Meal duration recorded by researcher and food intake divided by meal duration (kcal/minute)<br>Intervention (A), but not (B) differed significantly (faster) to Control<br>**Eating rate (M ± SD)** | Hunger and fullness self-reported VAS multiple times after meal (0-100, AUC)<br>Intervention B significantly differed (larger AUC) to Control, but Intervention A did not differ to Control for hunger<br>**Hunger (M ± SD)** | N/A |
|  |  | (B) 'Low nutritional quality'<br>Food and drink ED = 0.95<br>Food ED = 1.40 |  | ≈Fat<br>Sat fat (NR)<br>≈Carbohydrate<br>>Sugars<br>Sodium (NR)<br>≈Protein<br><Fibre |  | Intervention (A): 67.7 ± 23.1<br>Intervention (B): 50.1 ± 15.1<br>Control: 54.1 ± 18.4 | Intervention (A) = 127.7 ± 83.0<br>Intervention (B) = 159.7 ± 94.2<br>Control = 98.4 ± 69.9<br>No significant difference between trial arms for fullness<br>**Fullness (M ± SD)**<br>Intervention (A) = 293.1 ± 89.2<br>Intervention (B) = 265.4 ± 114.3<br>Control = 278.2 ± 82.0 |  |
| Lasschuijt et al. (2023) | Not reported<br>Trial design ensures large difference | (A) Soft texture food ED = 1.0<br>(B) Hard texture food ED = 1.0 | (A) Soft texture food ED = 1.0<br>(B) Hard texture food ED = 1.0 | (A) vs. Control:<br>≈Fat<br>>Sat fat<br>≈Carbohydrate<br><Sugars<br>>Sodium<br><Protein<br><Fibre<br>(B) vs. Control:<br>>Fat<br>>Sat fat<br>≈Carbohydrate<br><Sugars<br>>Sodium<br><Protein<br>>Fibre | Self-reported liking (9-point likert) before meals<br>No consistent significant differences between intervention vs. control arms across meals<br>**Liking (M ± SE)**<br>Intervention (A): 7.1 ± 0.3<br>Intervention (B): 7.0 ± 0.3<br>Control (A): 6.7 ± 0.3<br>Control (B): 7.2 ± 0.3 | Meal duration recorded via video and food intake divided by meal duration (kcal/minute)<br>Significant overall difference (faster) between intervention vs. control arms driven by Intervention A vs. Control A only<br>**Eating rate (M ± SE)**<br>Intervention (A): 77.0 ± 5.0<br>Intervention (B): 38.0 ± 5.0<br>Control (A): 62.0 ± 5.0<br>Control (B): 38.0 ± 5.0 | Hunger and fullness self-reported VAS before meals (0-100)<br>No consistent significant differences between intervention vs. control arms across meals for hunger or fullness<br>**Hunger (M ± SE)**<br>Intervention (A): 64.7 ± 4.3<br>Intervention (B): 61.7 ± 4.3<br>Control (A): 63.0 ± 5.0<br>Control (B): 66.3 ± 4.3<br>**Fullness (M ± SE)**<br>Intervention (A): 17.3 ± 4.3<br>Intervention (B): 19.7 ± 4.3<br>Control (A): 17.3 ± 4.3<br>Control (B): 18.0 ± 4.3 | N/A |
| Teo et al. (2022) | Not reported<br>Trial design ensures large difference | (A) Soft texture food ED = 1.23<br>(B) Hard texture food ED = 1.55 | (A) Soft texture food ED = 1.13<br>(B) Hard texture food ED = 1.11 | Data not reported | Self-reported liking (VAS; 0-100) after tasting meal<br>**Liking (M ± SE)**<br>Intervention (A): 68.4 ± 2.4<br>Intervention (B): 75.0 ± 2.8<br>Control (A): 66.3 ± 2.7<br>Control (B): 35.8 ± 3.3 | Meal duration recorded via video and food intake divided by meal duration (kcal/minute)<br>Significant differences (faster) between both | Hunger and fullness self-reported VAS multiple times after meal (0-100, AUC)<br>No significant difference between trial arms for hunger<br>**Hunger (M ± SE)** | N/A |
|  |  |  |  |  | Intervention vs. control differences not reported | (A, B) intervention vs. control arms<br>**Eating rate (M ± SE)**<br>Intervention (A): 66.6 ± 3.0<br>Intervention (B): 43.0 ± 1.8<br>Control (A): 53.5 ± 2.0<br>Control (B): 29.7 ± 1.5 | Intervention (A): 4.8 ± 3.2<br>Intervention (B): 12.7 ± 9.3<br>Control (A): 12.4 ± 8.7<br>Control (B): 14.6 ± 8.3<br>Intervention (A) vs. Control (A) did not significantly differ, but Intervention (B) vs. Control (B) significantly differed (larger AUC) for fullness<br>**Fullness (M ± SE)**<br>Intervention (A): 4243.7 ± 264.2<br>Intervention (B): 4072.5 ± 289.7<br>Control (A): 3661.8 ± 304.9<br>Control (B): 2913.5 ± 294.0 |  |

Abbreviations: ED = energy density, UPF = ultra-processed food. LPF = less processed food, AUC = area under the curve, VAS = visual analogue scale, > indicates UPF condition is at least 10% higher than LPF condition for nutrient, < indicates UPF condition is at least 10% lower than LPF condition for nutrient, ≈ indicates conditions are similar for nutrient (within 9.9%).

**Table 4.**
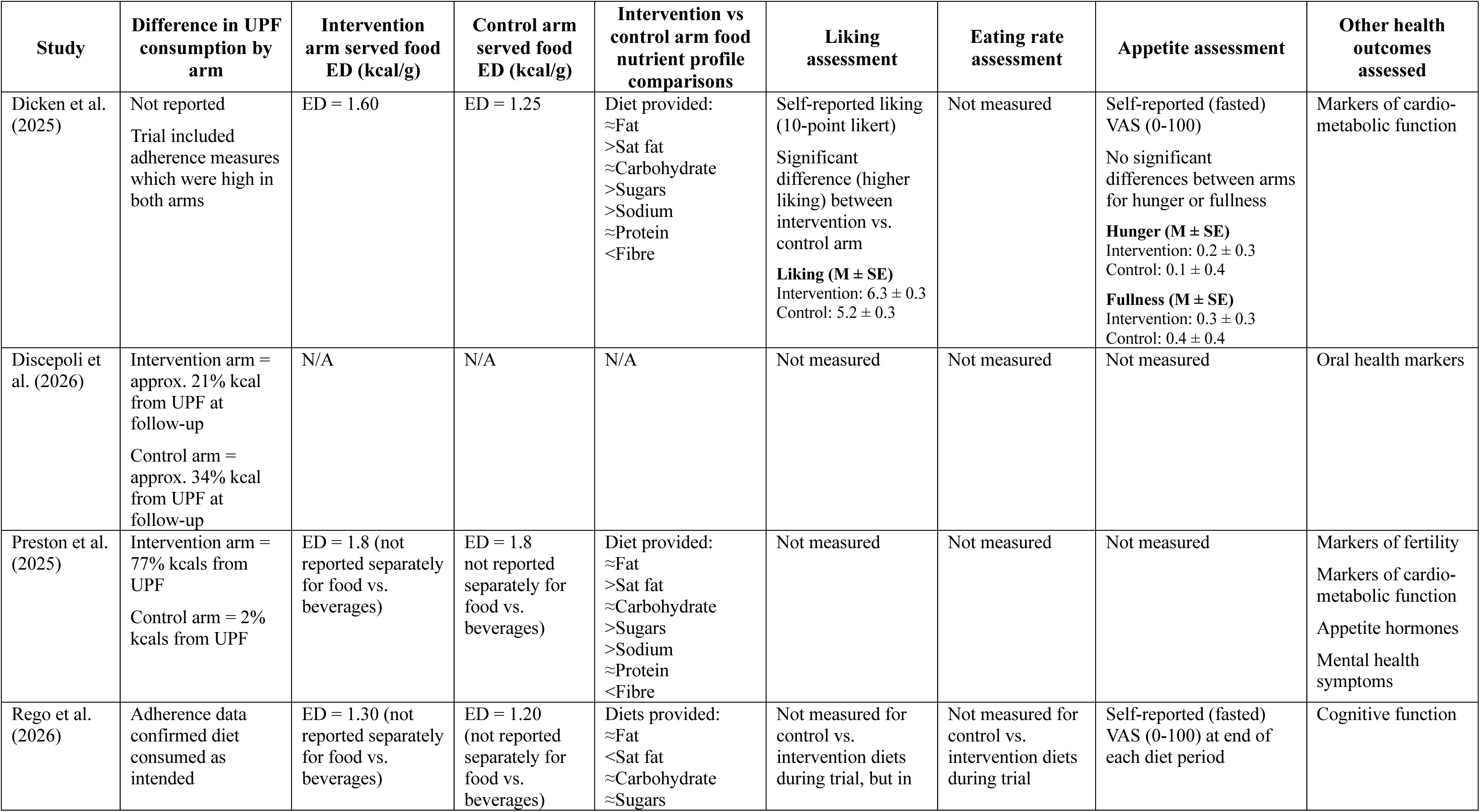
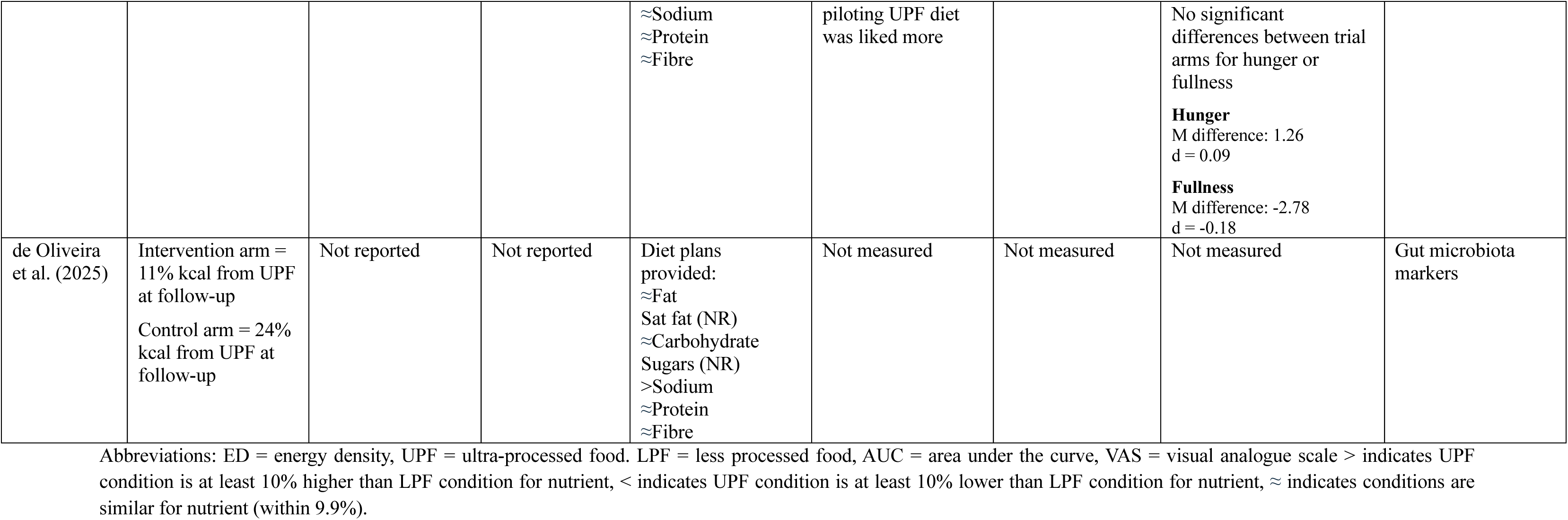
Additional methodological and secondary outcome information for included real-world studies.

### Primary outcomes Energy intake

Seven studies (Dicken et al., 2025; Discepoli et al., 2026; Hall et al., 2019; Hamano et al., 2024; Larcom et al., 2026; Lasschuijt et al., 2023; Teo et al., 2022) examined energy intake as an outcome and due to some studies comparing multiple forms of UPF in their design (e.g., soft and hard textured UPFs vs. soft and hard textured LPF), a total of 10 effects were included in meta-analysis. Energy intake was assessed at single meals, across a single day, and between 1 to 16 weeks. With all studies included, the overall pooled effect on energy intake was SMD = 0.34 ([95% CI: -0.03 to 0.70]; z = 1.74, p = 0.068, I^2^ = 88%, tau^2^ = 0.28], see Figure 2. Prediction intervals (95%) were -0.77 to 1.48, suggesting that 95% of effects from new studies would be expected to fall between these intervals. For laboratory studies (N = 5, k = 8) the pooled effect was SMD = 0.34 ([95% CI: -0.12 to 0.81]) (Hall et al., 2019; Hamano et al., 2024; Larcom et al., 2026; Lasschuijt et al., 2023; Teo et al., 2022) and for real world studies the pooled effect was similar (N = 2, k = 2: SMD = 0.39 [95% CI: 0.13 to 0.65]) (Dicken et al., 2025; Discepoli et al., 2026) (Figure 2).

**Figure 2.**
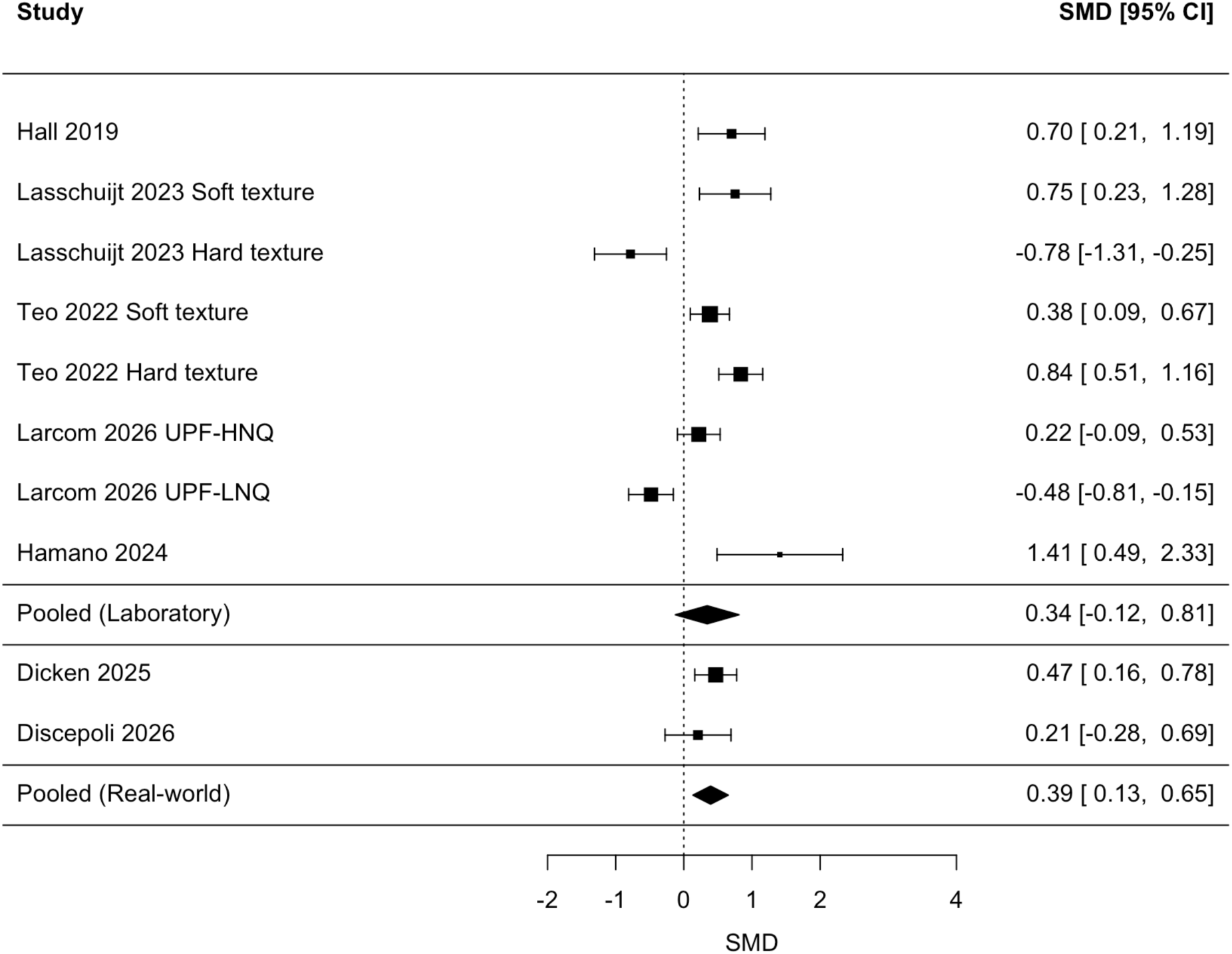
Forest plot for energy intake primary analysis, including all eligible studies. Positive SMD (standardised mean difference) indicates UPF trial arm had a greater energy intake than the comparison arm, negative SMD indicates the opposite. (Larcom 2026: UPF-HNQ: ultra-processed food high nutritional quality and UPF-LNQ: ultra-processed food low nutritional quality).

Statistical significance of the primary model varied dependent on inclusion/exclusion of some trials. Removal of the one outlying effect size (SMD = -0.78: hard texture (Lasschuijt et al., 2023)) increased the pooled effect size and made the overall model statistically significant (SMD = 0.47 [95% CI: 0.14 to 0.80]; z = 2.74, p = 0.005, I^2^ = 82.7%). Removal of one study (Discepoli et al., 2026) rated as being of poor methodological quality (see risk of bias section below) did not have a considerable influence on the pooled effect (SMD = 0.35 [95% CI: -0.05 to 0.76]; z = 1.70, p = 0.087, I^2^ = 89.9%). Removal of the largest positive effect size (Teo et al., 2022) reduced the pooled effect (SMD = 0.26 [95% CI: -0.09 to 0.61]; z = 1.45, p = 0.146, I^2^ = 87.4%). The funnel plot (all effect sizes) is shown in the supplementary materials and visual inspection was indicative of potential publication bias. Consistent with this, Trim and Fill analysis demonstrated that inclusion of two effects (small and negative) would lead to a reduction in the overall effect (SMD = 0.18 [95% CI: -0.19 to 0.56], p = 0.335) in the overall model, but due to the small number of effect sizes included evidence for publication bias should be interpreted with caution.

For descriptive purposes, meta-analysis of raw kcal differences limited to five studies (Dicken et al., 2025; Discepoli et al., 2026; Hall et al., 2019; Hamano et al., 2024; Lasschuijt et al., 2023) which measured daily energy intake (N = 5, k = 6) indicated a statistically significant +307.19 [95% CI: 3.29 to 611.10] kcal increase to daily energy intake in the UPF vs. LPF trial arms.

### Body weight

Five studies (de Oliveira et al., 2025; Dicken et al., 2025; Hall et al., 2019; Hamano et al., 2024; Preston et al., 2025) examined change in body weight as an outcome and due to one study examining two UPF conditions, a total of 6 effects were included in meta-analysis. Change in body weight was examined across 1 week to 6 months. The pooled effect of weight change was significant (SMD = 0.65 [95% CI: 0.38 to 0.94]; z = 4.55, p < 0.001, I^2^ = 40.9%, tau^2^ = 0.039: see Figure 3). The 95% prediction intervals were 0.16 to 1.16. There were no outlying effect sizes. Removal of the largest effect size and smallest effect size did not substantially influence results (largest removed: SMD = 0.60 [95% CI: 0.34 to 0.86], p < 0.001); smallest removed: SMD = 0.77 [95% CI: 0.52 to 1.03], p < 0.001). There were too few effects to reliably determine evidence for small study/publication bias via Trim and Fill procedure, see supplementary materials for funnel plot. For raw body weight change the pooled model suggested a significant increase in body weight between the UPF vs. comparator conditions (1.37kg [95% CI: 1.00 to 1.74], z = 7.20, p < 0.001).

**Figure 3.**
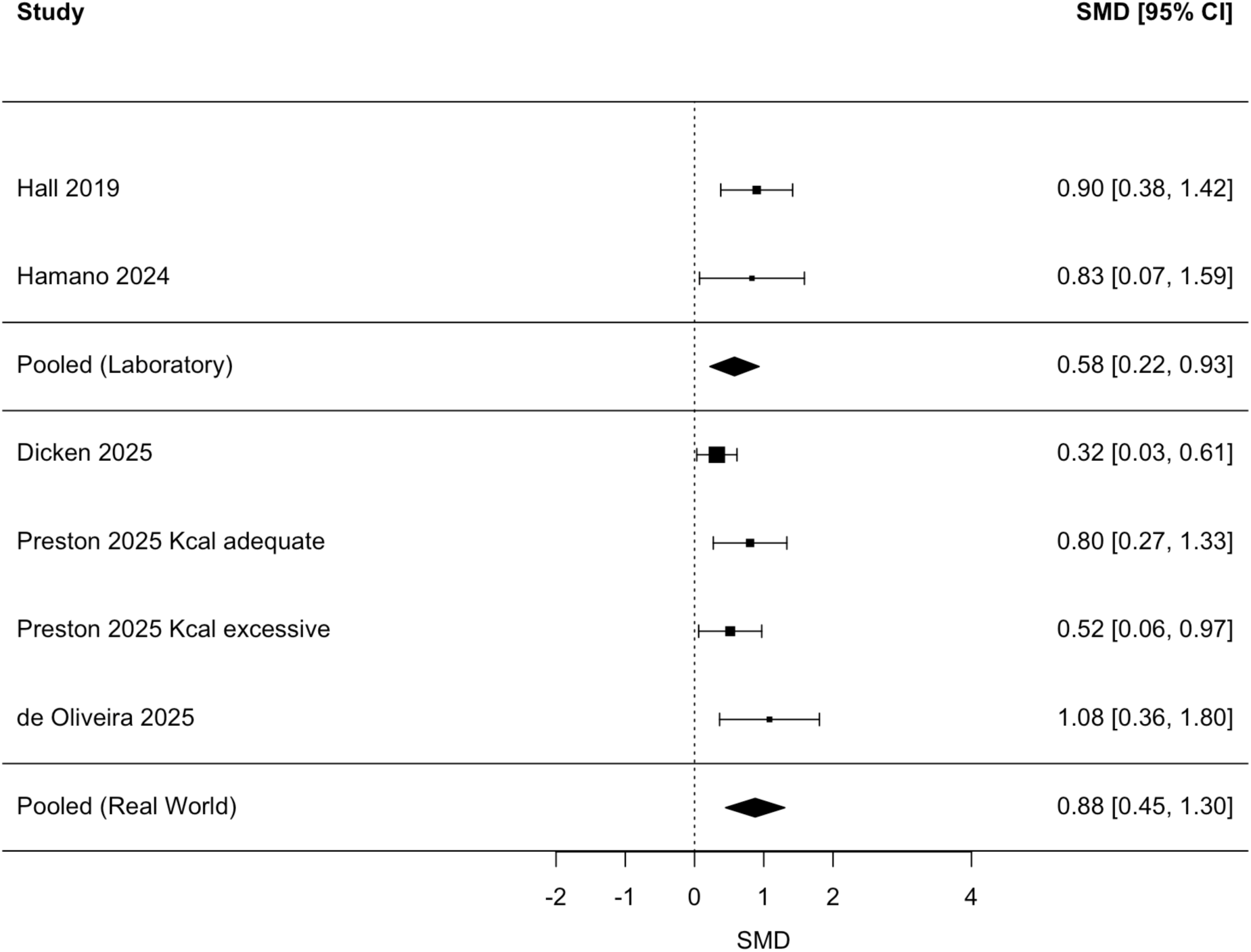
Forest plot for body weight changes primary analysis, including all eligible studies. Positive SMD (standardised mean difference) indicates UPF trial arm gained more weight outcome than the comparison arm, negative SMD indicates the opposite.

### Secondary outcomes

Studies measured self-reported liking/palatability of foods (N = 5, k = 7), researcher measured eating rate (N = 5, k = 8), self-reported hunger (N = 6, k = 9), and fullness (N = 6, k = 9) across trial arms. There was evidence of a significantly faster eating rate for UPFs vs. LPF, but no significance difference between trial arms for palatability, hunger or fullness.

For palatability, the pooled effect was SMD = 0.38 [95% CI: -0.06 to 0.81], z = 1.68, p = 0.092, I^2^ = 91%, tau^2^ = 0.36) (Dicken et al., 2025; Hall et al., 2019; Larcom et al., 2026; Lasschuijt et al., 2023; Teo et al., 2022). For eating rate, the pooled effect was SMD = 0.96 [95% CI: 0.12 to 1.80], z = 2.23, p = 0.026, I^2^ = 95%, tau^2^ = 0.636) (Hall et al., 2019; Hamano et al., 2024; Larcom et al., 2026; Lasschuijt et al., 2023; Teo et al., 2022) indicative of higher eating rate in the UPF trial arms. We therefore used meta-regression to examine if difference in eating rate between UPF vs. LPF arms in trial was associated with between arms differences in energy intake. There was a significant association between eating rate and energy intake (B = 0.070 [95% CI: 0.098 to 0.042], p < 0.001) (Figure 5); whereby, a larger difference in eating rate between UPF and LPF trial arms was predictive of a larger difference in energy intake between trial arms. For hunger, the pooled effect was SMD = 0.07 [95% CI: -0.16 to 0.30], z = 0.60, p = 0.547, I^2^ = 61%, tau^2^ = 0.058) (Dicken et al., 2025; Hall et al., 2019; Hamano et al., 2024; Larcom et al., 2026; Lasschuijt et al., 2023; Teo et al., 2022). For fullness, the pooled effect was SMD = 0.13 [95% CI: -0.05 to 0.31], z = 1.39, p = 0.162, I^2^ = 42%, tau^2^ = 0.024) (Dicken et al., 2025; Hall et al., 2019; Hamano et al., 2024; Larcom et al., 2026; Lasschuijt et al., 2023; Teo et al., 2022).

**Figure 4.**
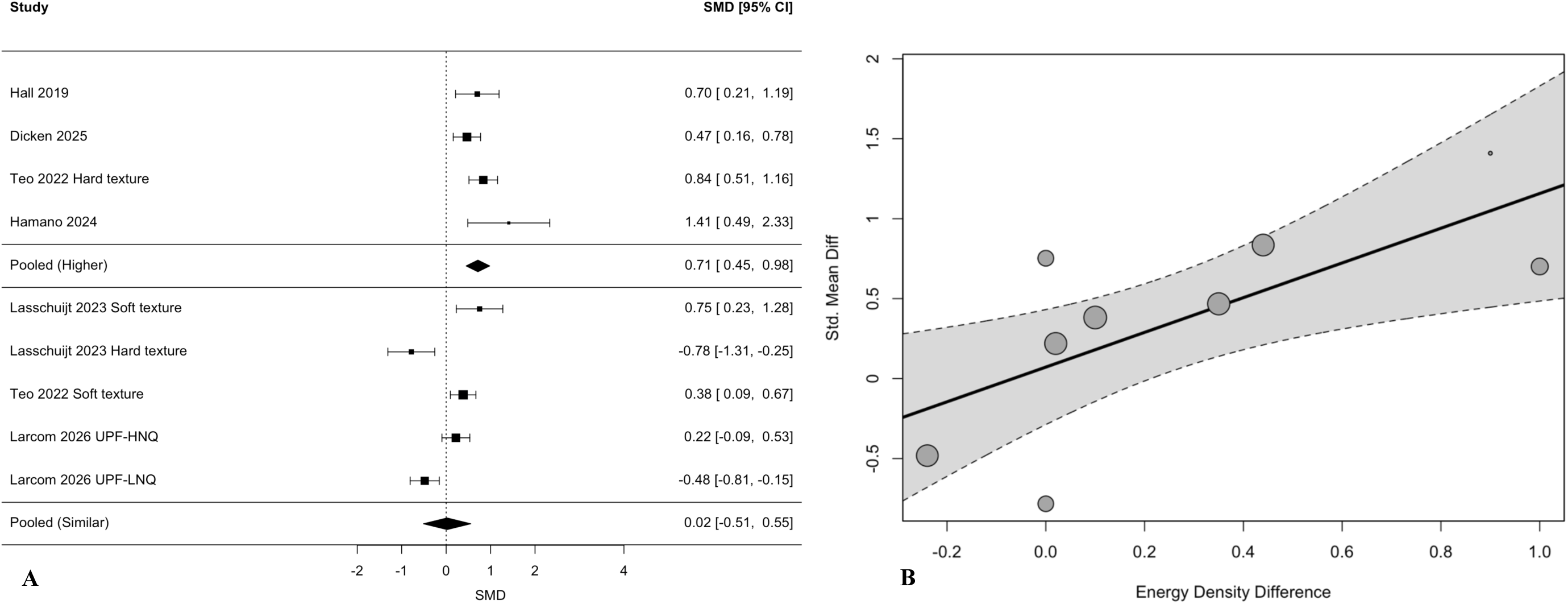
Food provision studies by **(A)** examining effects on energy intake by energy density categorisation (Larcom 2026: UPF-HNQ: ultra-processed food high nutritional quality and UPF-LNQ: ultra-processed food low nutritional quality). Positive SMD (standardised mean difference) indicates UPF trial arm has a higher score on the outcome (energy intake) than other arm, negative SMD indicates the opposite. Higher indicates trial arm serving predominantly UPF had a higher energy density than other arm. Similar indicates trial arm serving predominantly UPF had a similar or slightly lower energy density than other arm; and **(B)** association between trial arms for energy density and energy intake differences.

**Figure 5.**
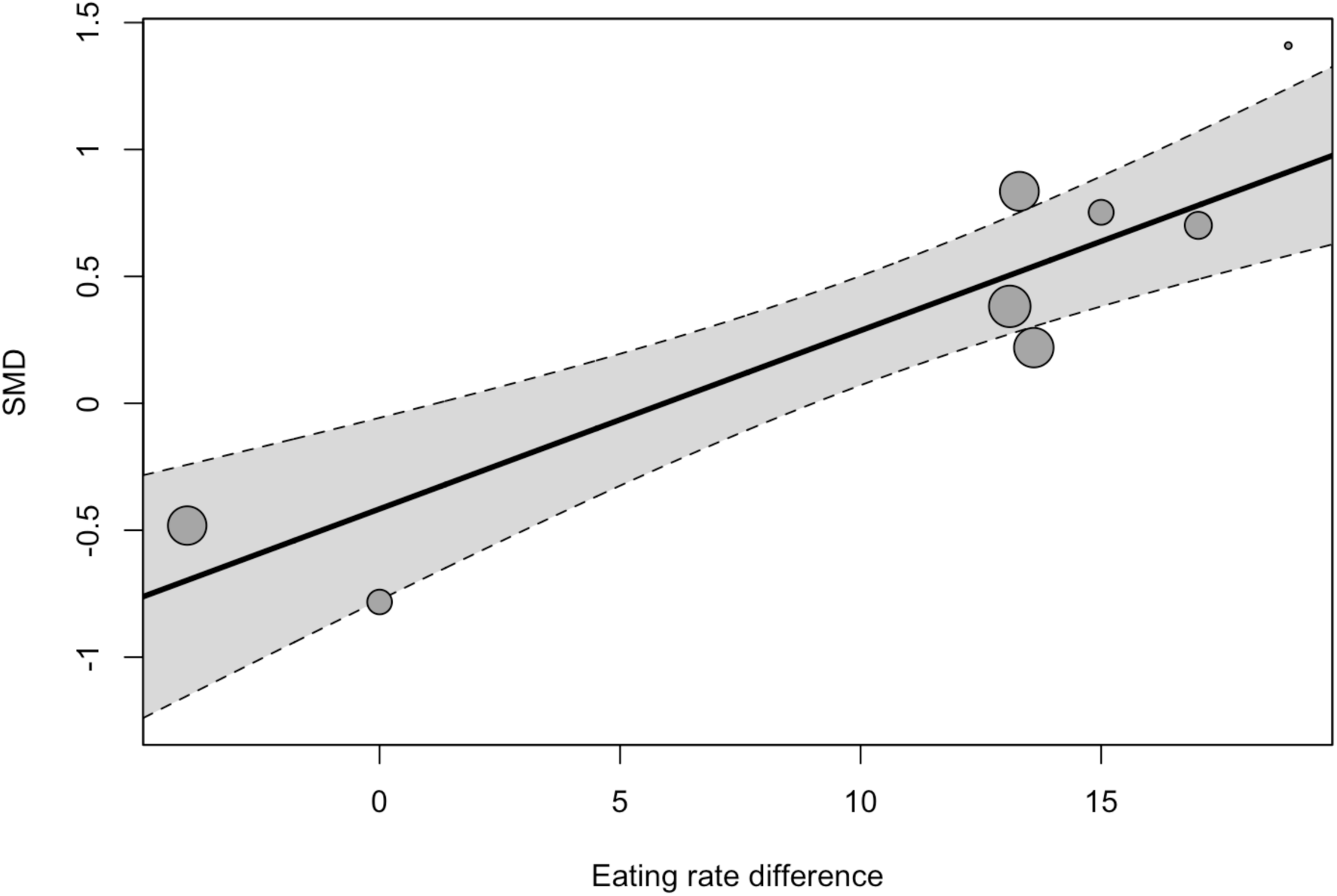
Association between trial arms for eating rate and energy intake differences (SMD = standardised mean difference).

Three studies (Larcom et al., 2026; Rego et al., 2026; Teo et al., 2022) examined subsequent energy intake (i.e., energy intake measured after provision of UPF vs. LPF) in the form of energy intake for the remainder of a day after being provided with a UPF vs. LPF breakfast or after 14 days of UPF vs. LPF diet provision. In all three studies there were no significant differences between trial arms for subsequent energy intake.

### Evidence for effect of processing independent to nutrient profile

Studies did not match UPF vs. LPF diets for nutrients profile, with UPF diet arms typically higher in saturated fat, salt and sugar, and lower in fibre, resulting in higher average dietary energy density. See Table 3-4. For energy intake outcome studies, energy density of UPF diets were markedly higher than corresponding LPF (intervention) diets (>10%) for k = 4 comparisons, compared to k = 5 comparisons in which energy density of UPFs was similar or lower than LPF. Examined as a formal moderator, dietary energy density difference significantly predicted energy intake (X^2^(1) = 4.86, p = 0.028). For studies which served participants UPFs that were more energy dense than the LPF diet, the pooled effect was SMD = 0.71 [95% CI: 0.45 to 0.98]; z = 5.29, p < 0.001 (Dicken et al., 2025; Hall et al., 2019; Hamano et al., 2024; Teo et al., 2022 (hard texture)) (Figure 4). For studies which served participants UPFs that had a similar or lower energy density than the LPF diet, there was no effect of UPF on energy intake (SMD = 0.02 [95% CI: - 0.51 to 0.55]; z = 0.079, p = 0.937) (Teo et al., 2022 (soft texture); Larcom et al., 2026 (UPF HNQ+LNQ); Lasschuijt et al., 2023 (soft+hard texture)) (Figure 4). Consistent with this, there was a strong significant association between individual study effect sizes and the size of energy density difference between the UPF and LPF conditions (B = 1.09 [95% CI: 0.29 to 1.88], z = 2.67, p = 0.008: see Figure 4). There were too few studies and variations in energy density between trial arms for body weight change outcome studies to formally examine moderation by energy density difference. All relevant forest plots are in supplementary materials.

### Risk of bias

For examination of the overall pooled effects of UPF on energy intake and weight gain (objective one), risk of bias tended to be low with eight of ten studies categorised as being ‘Good’ (de Oliveira et al., 2025; Hall et al., 2019; Hamano et al., 2024; Larcom et al., 2026; Lasschuijt et al., 2023; Preston et al., 2025; Rego et al., 2026; Teo et al., 2022). The remaining two studies (Dicken et al., 2025; Discepoli et al., 2026) were both real-world trials rated as ‘Neutral’ and ‘Poor’. See supplementary materials for individual study level risk of bias scores. We considered risk of bias to be very high when assessing evidence for effects of processing independent to nutritional profile of food (objective two). This was because in all studies included there was either a lack of matching for key nutrients between UPF compared with LPF provided to participants and/or studies did not report complete information on nutritional composition of diets provided to participants. See Tables 3-4.

### Certainty of evidence

We rated certainty of evidence for the pooled effects of UPF compared with LPF diets on energy intake to be low and change in body weight to be moderate to low because of the limited studies available, some evidence of potential publication bias and analyses indicating effects on energy intake were dependent on inclusion/exclusion of individual trials. These observations reduce certainty in whether the pooled effects observed in the present meta-analyses are reflective of the impact UPF would reliably have on studied outcomes. We rated evidential certainty as being very low for any effects of food processing on studied outcomes operating independently to nutritional profile, due to the aforementioned limitations in nutritional matching of diets in reviewed trials and energy density sub-group findings.

## 4. Discussion

Pooled analysis of ten RCTs compared the effect of UPF with LPF on energy intake and body weight. This provided new insight by synthesising all currently available evidence and reviewed evidential certainty finding: (i) mixed evidence on the impact UPF has on energy intake; therefore, potentially challenging common assumptions that UPF increases energy intake and (ii) more consistent evidence for impact on increased weight gain, albeit from a smaller subset of trials. Due to the nutritional profile of foods typically used in RCTs differing between trial arms, there was no convincing evidence that effects on energy intake and weight gain observed were independent to nutrient profile of UPFs. Instead, sub-group analyses were consistent with UPF effects on energy intake in trials to date potentially being explained by dietary energy density differences between UPFs and LPF used in RCTs to date.

Secondary outcome analyses from a smaller sub-set of studies revealed no effects of UPF vs. LPF on reported palatability of diet, appetite or subsequent energy intake, which is not consistent with suggestions that UPF may promote weight gain due to its hyper-palatability and/or lack of satiating effects (Juul et al., 2025). UPFs had a faster eating rate than LPF (on average) and this may be explained by foods higher in energy density promoting faster eating rates (kcal/min) and/or texture and food matrix differences between UPF compared with LPF used in RCTs (Forde et al., 2020). Consistent with this, we found evidence that differences in eating rate between UPF and LPF trial arms were associated with effects on energy intake, whereby UPF arms with a faster eating rate tended to have a higher energy intake.

A range of other health markers were examined in reviewed RCTs, including cardiometabolic, fertility and dental outcomes. As discussed, RCTs examined tended not to match UPF vs. LPF for key nutrients (e.g., fibre, fat, sugar) and dietary energy density. Many of these factors are already known to affect appetite, as well as cardiometabolic, fertility and dental health outcomes (Funtikova et al., 2015; Moores et al., 2022; Panth et al., 2018). To enable conclusions to be drawn on any effects of ultra-processing independent to nutrient profile, future RCTs examining such health outcomes will need to more closely match diets, including non-beverage energy, consumed by participants. Recent observational studies suggest that it may be specific sub-groups of UPF (such as animal-based products and sugar-sweetened beverages) that increase risk of adverse health outcomes (Cordova et al., 2023). However, we found that RCTs did not consistently provide information on processing level of LPF provided to participants (e.g., ‘minimally processed’ vs. ‘processed’) or food characteristics (e.g., ingredients, presence of specific additives or processing techniques). More accurate characterisation of UPF vs. LPF provided to participants in future trials will now be valuable.

Due to the limited number of RCTs conducted to date, conclusions drawn from the present analyses are made tentatively and findings should be interpreted cautiously. Yet, the present synthesis of current evidence is timely due to there being significant public and policy interest in UPFs. Furthermore, our evidence synthesis and identification of current research gaps provides useful guidance to ensure future RCTs are well-designed and informative. Whilst a number of RCTs described UPF compared with LPF diets as being closely matched for nutritional profile, this was often not the case for the main source of energy in the diet, foods, as opposed to beverages. Future studies should therefore account for the contribution of different energy sources in the diet when matching nutritional profile. Future studies examining effects on health outcomes of changing UPF consumption via dietary advice will also benefit from using active control groups, as opposed to wait list controls due to inherent biases associated with the latter (Cunningham et al., 2013) which may lead to erroneous conclusions about the causal impact of UPF on health outcomes (Waters et al., 2012). See Table 5 for a summary of recommendations for future RCTs.

**Table 5.** Recommendations for future randomised controlled trials.

| Recommendation | Rationale |
| --- | --- |
| Studies designed to match diets for nutrient profile should account for size of contribution study foods and/or drinks will likely make towards total energy intake | Matching of nutrient profile by overall energy provided in diet only potentially results in imbalances of nutrient profile for major sources of energy (e.g., main meal foods) and therefore total nutrient intake |
| Studies designed to match diets for nutrient profile should consider whether the degree of difference between diets for nutrients (e.g., <5% difference) is acceptable based on hypotheses | Relatively small differences in individual nutrients can additively contribute to meaningful differences in overall nutritional profile (e.g., differences in energy density) |
| Studies should routinely measure and account (where appropriate) for key factors known to influence energy intake which may co-vary by diets provided | Not measuring or accounting for potential diet arm differences in known drivers of energy intake (e.g., food palatability, texture) does not allow their contribution to energy intake to be considered |
| Studies examining effects of UPF should routinely describe the proportion of provided and consumed diet in each trial arm from different processing categories (% minimally processed, processed, UPF) | Not reporting processing level of diet consistently limits interpretation of findings and potential for future research syntheses (e.g., extent to which between trial arm differences in outcome variables is dependent on Nova category UPF is compared to) |
| Studies should provide detailed diet ingredient information and where possible, description of processing techniques of study foods | Not reporting ingredient and/or food processing information limits potential for future research syntheses considering ingredient and processing techniques |
| Studies should measure participant habitual consumption of UPFs and study foods to aid interpretation of how provision of study foods alters usual diet (e.g., increasing vs decreasing UPF/minimally processed/processed diet share) | Not measuring habitual consumption may prevent inferences being made about the impact that different provisions of food (e.g., effect of increasing, decreasing or maintaining habitual UPF vs. minimally processed food) have on outcome variables |
| Where possible, studies should actively blind participants to study aims (e.g., effect of UPF on energy intake and/or body weight), as opposed to explicitly informing participants of study aims | Participants made aware of study aims can change their behaviour, introducing bias which may explain trial arm differences in outcome variables |
| Studies changing UPF consumption through dietary guidance (as opposed to food provision) should use active control groups with appropriate intervention intensity matching | Active control groups separate out the specific effects of an intervention from being actively enrolled in a trial and motivated to improve health, whereas waitlist controls cannot achieve this, which introduces bias and may explain trial arm differences in behavioural outcome |

Reviewed studies had a number of strengths (e.g., objective measurement of outcome variables, relatively high compliance and low drop out) and tended to not be high in risk of bias. Limitations of our review include the small number of RCTs available to synthesise as well as incorporating different study designs and participant groups (healthy, overweight, obese) to capture all available data. This reduced certainty in evidence due to high heterogeneity and wide prediction intervals; therefore, suggests that results of future meta-analyses could feasibly change when a larger body of evidence becomes available. Body weight change tended to be examined over relatively short time periods and this varied between RCTs, as was the case for energy intake. RCTs reviewed also tended to compare the effect of UPF to foods predominantly from Nova category 1 (‘minimally processed’) or did not report this information, so generalisability of results to comparison with other Nova categories is unclear.

These limitations aside, the present analysis is the first to quantitatively synthesise available evidence from RCTs of the effects UPF compared with LPF has on energy intake and body weight. RCTs to date are suggestive that UPFs may increase energy intake and body weight, but evidential certainty is low. At present there is no convincing evidence that ultra-processing of food has an effect on energy intake or body weight independent to nutritional profile.

## 5. Conclusion

RCTs are suggestive that UPFs may increase energy intake and body weight, although results may be explained by energy density of foods used in RCTs to date. Further research is needed to understand whether level of ultra-processing impacts health outcomes independent to nutrient profile.

## Author Contributions

ER drafted the manuscript. All authors contributed to protocol development and/or research methods, in addition to reviewing the manuscript prior to submission.

## Data Availability

All data produced are available online at Open Science Framework (https://osf.io/sdrcz).

## Acknowledgements/Funding

ER is funded by the National Institute for Health and Care Research (NIHR) Oxford Health Biomedical Research Centre (BRC), the MRC and the ESRC. VN is supported by an ESRC research grant and the NIHR Oxford Health BRC.

## Data Sharing

Data described in the manuscript will be publicly and freely available without restriction at the Open Science Framework (https://osf.io/sdrcz).

