## Supplementary for "A systematic review and meta-analysis of randomised controlled trials examining the effect of ultra-processed food on energy intake and weight gain"

### Supplementary Materials

#### Search strategy information

**Table S1.** Search strategies used in electronic databases.

| Database | Search strategy | Fields searched | Limits/filters applied | Records identified | Notes/source |
|---|---|---|---|---|---|
| Cochrane Library | (“ultra-processed food*” OR “ultra processed food*” OR “UPF” OR NOVA) AND (“energy intake” OR “calori* intake” OR “diet” OR “dietary intake” OR weight OR BMI OR “body mass index” OR obesity) | Title, Abstract, Keywords | Trials only<br><br>2009 - 2026 | 349 | https://www.cochranelibrary.com/search<br><br>Conducted 13/11/2025. |
| Scopus | TITLE-ABS-KEY (“ultra-processed food*” OR “ultra processed food*” OR “UPF” OR NOVA) AND TITLE-ABS-KEY (“energy intake” OR “calori* intake” OR “diet” OR “dietary intake” OR weight OR BMI OR “body mass index” OR obesity) | Title, Abstract, Keywords | Document type: Article (ar);<br><br>2009 - 2026 | 3322 | https://www.scopus.com/<br><br>Conducted 13/11/2025. |
| PubMed | (“ultra-processed food”[tw] OR “ultra processed food”[tw] OR “ultra-processed foods”[tw] OR “ultra processed foods”[tw] OR UPF[tw] OR NOVA[tw]) AND (“energy intake”[tw] OR “calori* intake”[tw] OR “diet”[tw] OR “dietary intake”[tw] OR weight[tw] OR BMI[tw] OR “body mass index”[tw] OR obesity[tw]) | Title, Abstract, Keywords, Indexing text | Filters: Adaptive Clinical Trial; Clinical Trial (Phases I - IV); Randomized Controlled Trial<br><br>2009 - 2026 | 87 | https://pubmed.ncbi.nlm.nih.gov/advanced/<br><br>Conducted 13/11/2025. |

**Reasons for exclusion of non-eligible studies**

**Table S2.** List of excluded studies with reasons for exclusion, note studies can be excluded for multiple reasons and table below provides primary reason chosen during screening.

| Study | Main reason for exclusion |
|---|---|
| Abar et al. (2025) | Wrong outcome |
| Baratto et al. (2025) | Wrong outcome |
| Baroni et al. (2024) | Wrong study design |
| Barros et al. (2025) | Wrong outcome |
| Brandão et al. (2024) | Wrong outcome |
| Brichacek et al. (2024) | Wrong outcome |
| Byker-Shanks et al. (2022) | Wrong outcome |
| Capra et al. (2024) | Wrong outcome |
| Cecchini et al. (2025) | Wrong study design |
| Chen et al. (2022) | Wrong outcome |
| Chun (2025) | Wrong publication type |
| Cortes et al. (2025) | Wrong study design |
| Cortes et al. (2023) | Wrong outcome |
| Crivellenti et al. (2025) | Wrong outcome |
| de Vargas et al. (2024) | Wrong publication type |
| Dicken et al. (2025a) | Wrong publication type |
| Dinu et al. (2024) | Wrong publication type |
| Dioneda et al. (2020) | Wrong outcome |
| Dionne et al. (2025) | Wrong outcome |
| Dionne et al. (2024) | Wrong outcome |
| Dos Santos Moraes et al. (2024) | Wrong outcome |
| Field et al. (2022) | Wrong outcome |
| Galdino-Silva et al. (2024) | Wrong outcome |
| Godsey et al. (2025) | Wrong study design |
| Godsey et al. (2024) | Wrong study design |
| Graciliano et al. (2025) | Wrong outcome |
| Green et al. (2024) | Wrong outcome |
| Hagerman et al. (2024) | Wrong study design |
| Heuchan et al. (2025) | Wrong study design |
| Iglésies-Grau et al. (2023) | Wrong outcome |
| Iglésies-Grau et al. (2024) | Wrong study design |
| Jaime-Lara et al. (2023) | Secondary data from eligible study |
| Joseph et al. (2020) | Secondary data from eligible study |
| Larcom et al. (2025) | Wrong publication type |
| K Liu et al. (2025) | Wrong outcome |
| K. Liu et al. (2025) | Wrong study design |
| Lopes et al. (2025) | Wrong study design |
| NCT04280146 (2020) | Wrong publication type |
| NCT05319327 (2022) | Wrong study design |
| NCT05539222 (2022) | Wrong study design |
| NCT05550818 (2022) | Wrong publication type |

| | |
|---|---|
| NCT05658757 (2022) | Wrong study design |
| NCT06044285 (2023) | Trial in progress |
| NCT06356220 (2024) | Wrong study design |
| NCT06538831 (2024) | Wrong outcome |
| NCT06907862 (2025) | Wrong study design |
| NCT06920914 (2024) | Trial in progress |
| NCT07175701 (2025) | Wrong study design |
| NCT07213245 (2025) | Trial in progress |
| O'Connor et al. (2023) | Secondary data from eligible study |
| Poll et al. (2020) | Wrong study design |
| RBR-10gfr3fb (2025) | Wrong publication type |
| RBR-3dbxrc2 (2024) | Wrong study design |
| RBR-3p52g2p (2025) | Wrong study design |
| RBR-3q9vgk9 (2023) | Wrong study design |
| RBR-48kjwtj (2022) | Wrong study design |
| RBR-4mp6nvg (2025) | Wrong study design |
| RBR-4pdv53d (2023) | Wrong study design |
| RBR-56nsh92 (2023) | Wrong outcome |
| RBR-6zm7yvg (2025) | Wrong publication type |
| RBR-9crqgt (2019) | Wrong study design |
| Reynolds et al. (2023) | Wrong study design |
| Salazar et al. (2025) | Wrong study design |
| Santos et al. (2024) | Wrong study design |
| Sartorelli et al. (2020) | Secondary data from eligible study |
| Sartorelli et al. (2023) | Wrong outcome |
| Sciarrillo et al. (2023) | Wrong publication type |
| Sciarrillo et al. (2024) | Secondary data from eligible study |
| Seibold et al. (2024) | Wrong outcome |
| Sneed et al. (2025) | Wrong study design |
| Valmorbida et al. (2023) | Wrong study design |
| Walker et al. (2022) | Wrong study design |
| Yao et al. (2024) | Wrong study design |

#### Additional analysis information

*Effect size computation and analysis strategy*

Analyses were conducted using the 'metafor' package.

Effect sizes were calculated using the 'escalc' function on raw data (group means, standard deviations and Ns). For the between-subjects RCT we used the "SMD" measure to get the effect size. For comparative effect sizes from within-subjects / cross-over trials we used the "SMCC" measure. For within-subjects / cross-over trials that required the correlation for a measured outcome between UPF vs. LPF arms but did not report this, we requested this information from authors for primary outcomes (energy intake, weight gain). For secondary outcomes, this information was not routinely available and we therefore imputed the size of the correlation (r = 0.5) and conducted sensitivity analyses in which we varied the size of imputed correlation (0.3 – 0.7), finding that results of all secondary outcome analyses did not differ under different imputation conditions.

As multiple effect sizes could be computed from a single study (e.g., Larcom et al., 2026; Teo et al., 2022; Lasschuijt et al., 2023) we conducted multilevel meta-analysis by including a random intercept for study ID, via the 'rma.mv' function. These models were largely consistent with single level models, and we retained the pooled estimates from multilevel models throughout.

*Publication and influential case analysis*

We assessed funnel plot asymmetry visually and dependent on number of studies, use Egger's test and/or the Trim and Fill procedure, if appropriate (i.e., 10 or more studies for eligible outcome). Note, the Trim and Fill procedure is only compatible with single level models. We also identified outliers using the 'boxplot' function. We removed outliers as sensitivity analyses as well as removing both the largest and smallest effect size from the primary outcomes.

#### Risk of bias information for included studies

**Table S3.** Summary of Nutrition Quality Evaluation Strengthening Tools (NUQUEST) ratings for risk of bias.

| Type | Study | Selection of participants | Comparability of study groups | Ascertainment of outcomes | Nutrition-specific | Overall study rating |
|---|---|---|---|---|---|---|
| Laboratory Studies | Hall et al. (2019) | Good | Good | Good | Good | **Good** |
| | Hamano et al. (2024) | Good | Good | Good | Good | **Good** |
| | (Larcom et al., 2026) | Good | Good | Good | Good | **Good** |
| | (Teo et al., 2022) | Good | Good | Good | Good | **Good** |
| | Lasschuijt et al. (2023) | Good | Neutral | Good | Good | **Good** |
| Real World Studies | Preston et al. (2025) | Good | Neutral | Good | Good | **Good** |
| | Dicken et al. (2025b) | Good | Neutral | Neutral | Good | **Neutral**[1] |
| | Discepoli et al. (2026) | Good | Poor | Poor | Poor | **Poor**[2] |
| | de Oliveira et al. (2025) | Good | Good | Good | Neutral | **Good** |
| | Rego et al. (2026) | Neutral | Good | Good | Good | **Good** |

Criteria for risk of bias (RoB) in accordance with Kelly et al. (2022): (i) Good (+): almost all criteria met, there is little or no concern and low RoB; (ii) Neutral (0): most criteria met but there are some flaws with an associated concern and moderate RoB; and (iii) Poor (-): most or all criteria not met, there are significant flaws and high RoB.

Rationale for overall downgrade based on:

[1]Differences in meal presentation between arms (i.e., food packaging) and potential bias in self-reported energy intake (e.g., lack of information on exclusion of implausible values)

[2]Interventation groups were not closely matched (non-active comparison group) and measurement outcome for energy intake introduces significant bias (food frequency questionnaire, no information on plausibility of values).

**Supplementary analysis figures**

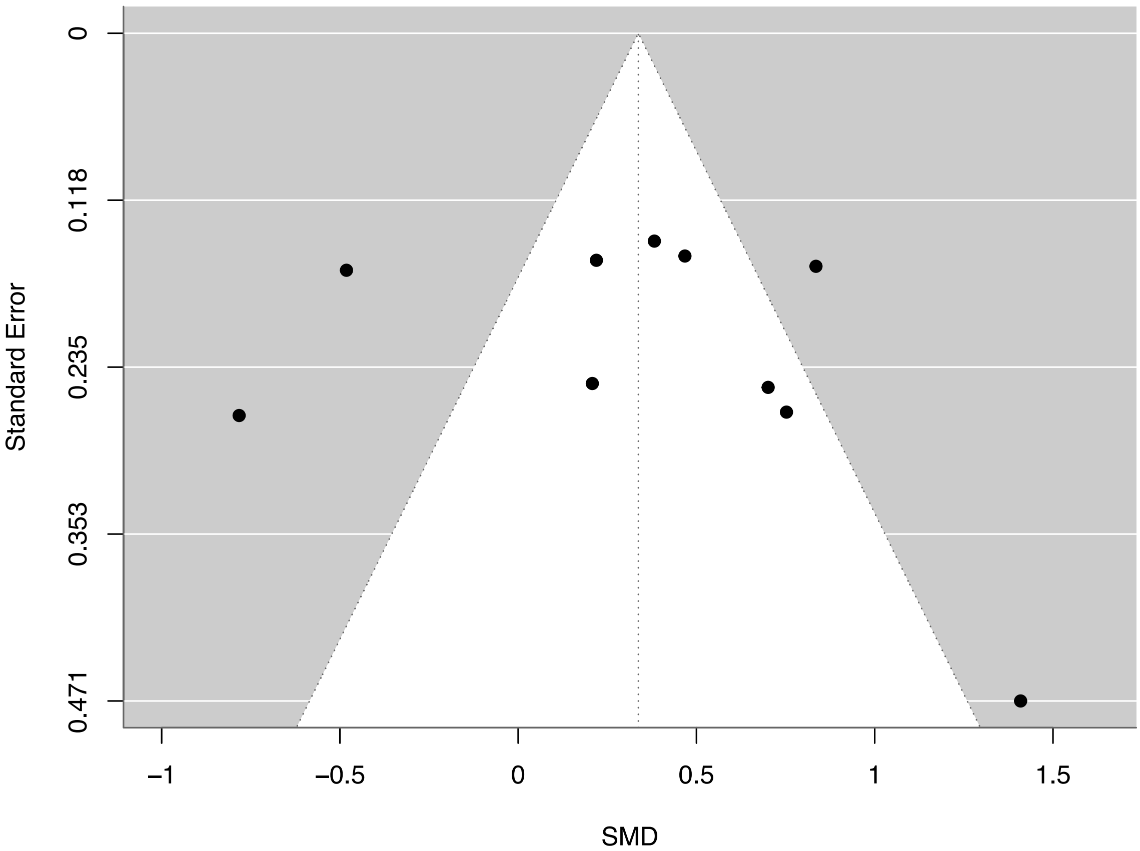


**Figure S1.** Funnel plot for energy intake studies (SMD: standardised mean difference).

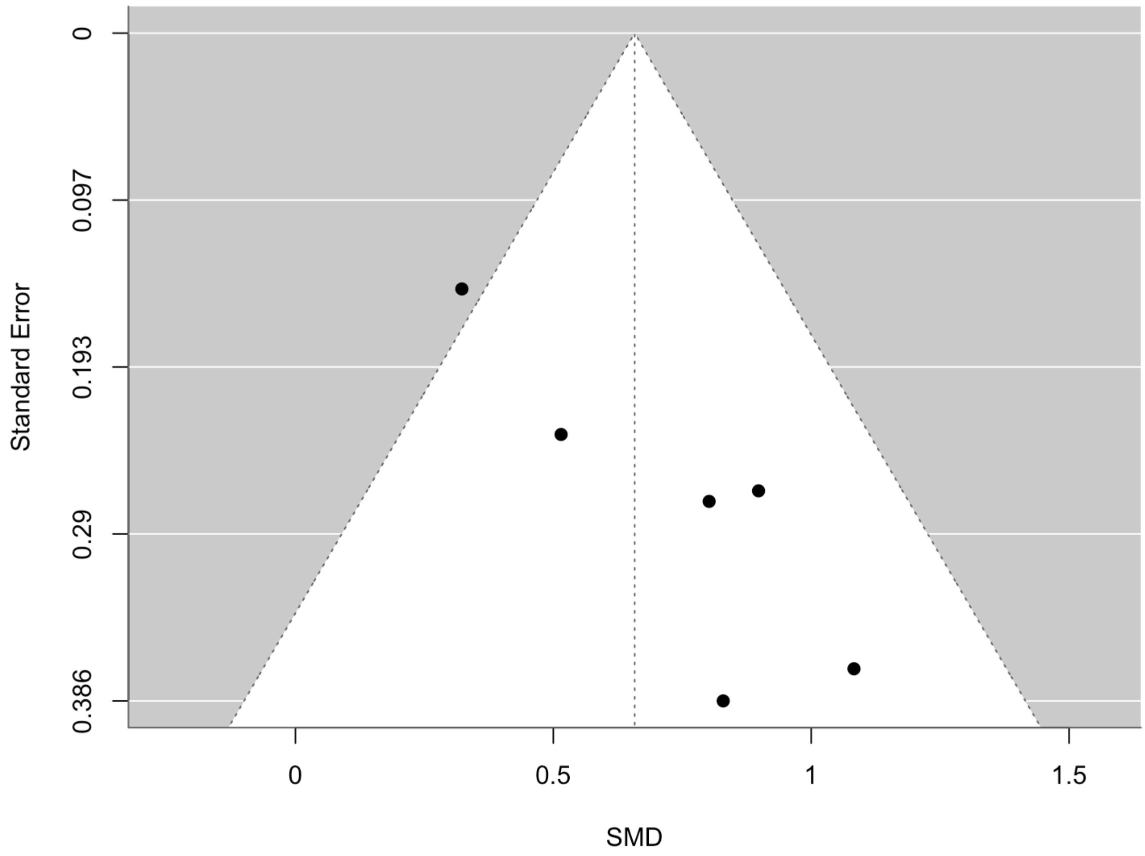


**Figure S2.** Funnel plot for weight change studies (SMD: standardised mean difference).

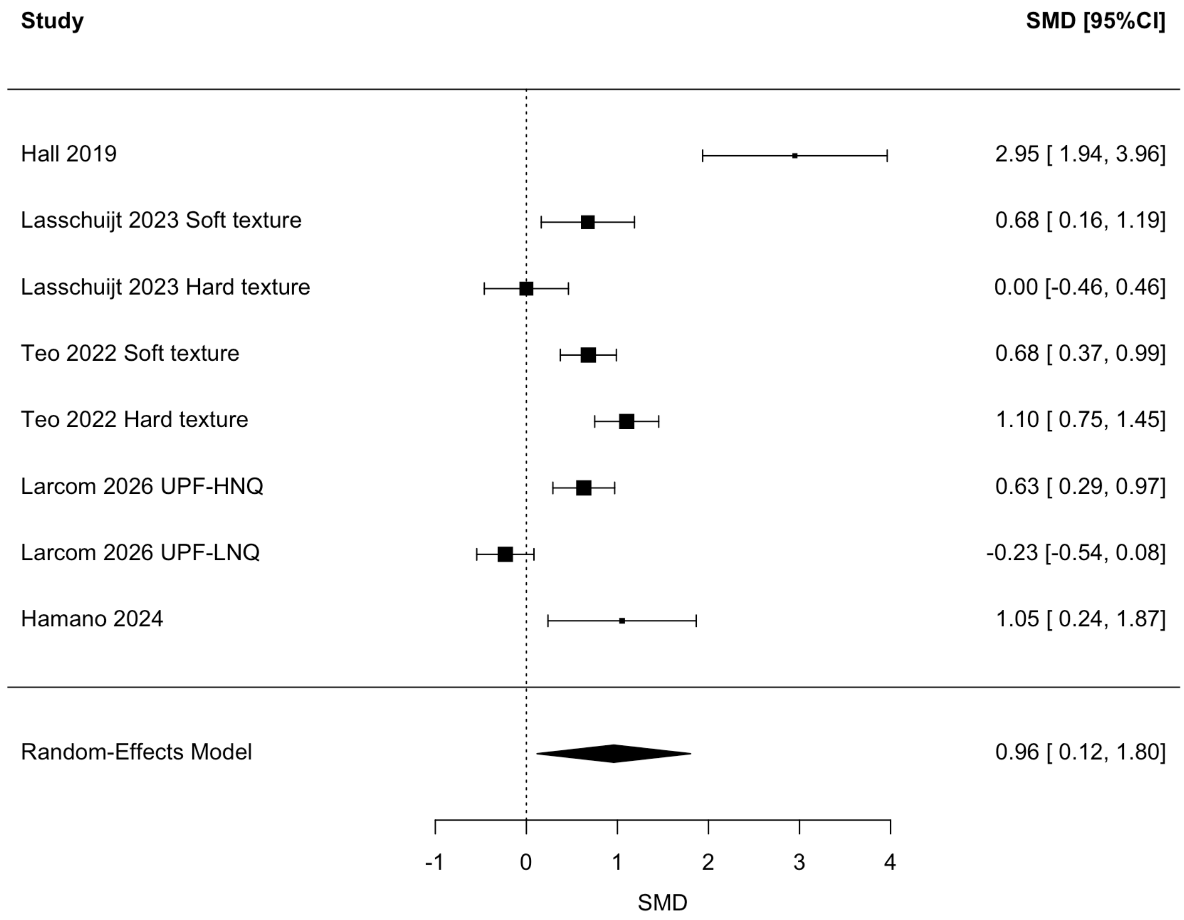


**Figure S3.** Eating rate meta-analysis (Larcom 2026: UPF-HNQ: ultra-processed food high nutritional quality and UPF-LNQ: ultra-processed food low nutritional quality). Positive SMD (standardised mean difference) indicates UPF trial arm has a higher score on the outcome (eating rate) than other arm, negative SMD indicates the opposite.

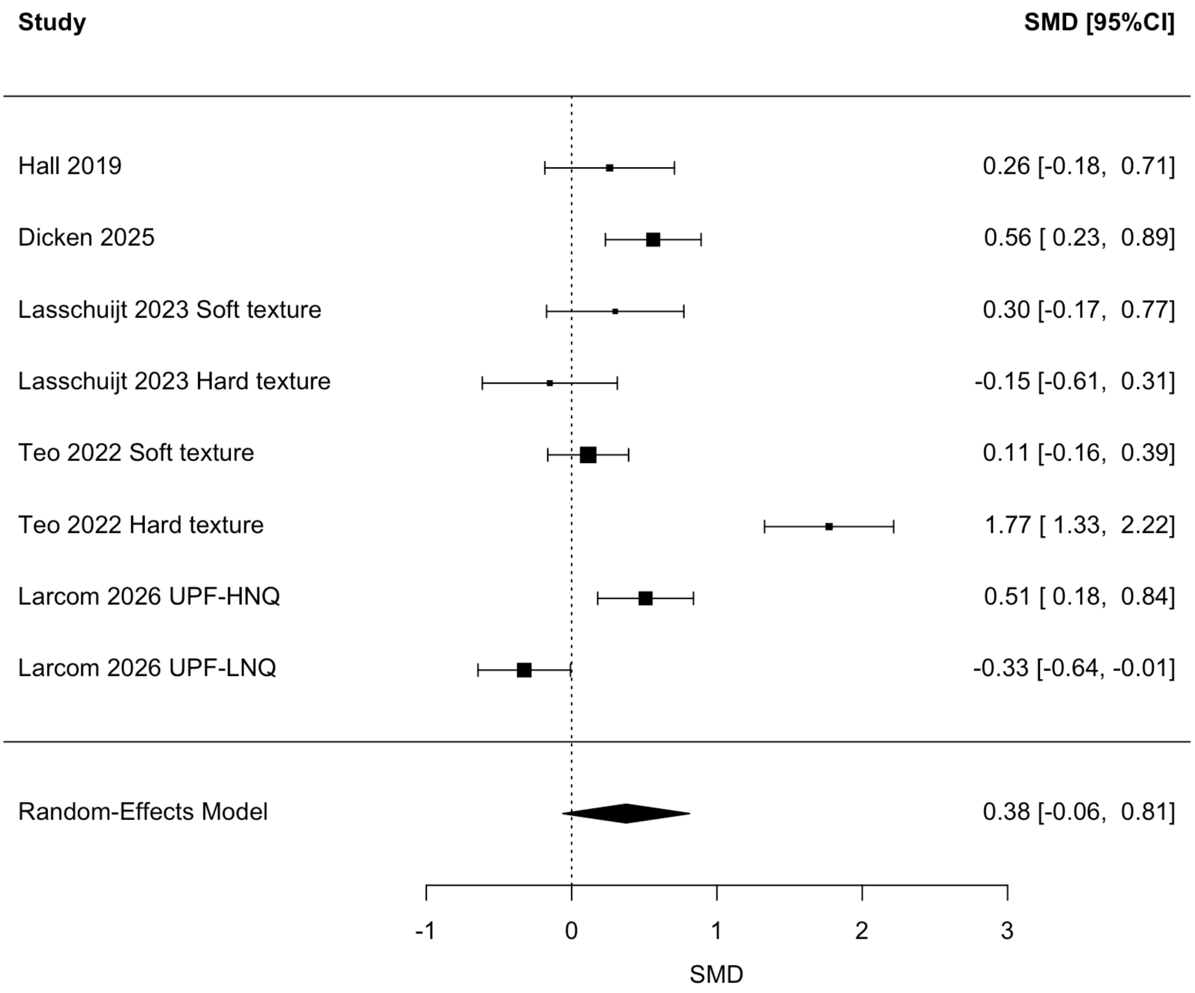


**Figure S4.** Palatability meta-analysis (Larcom 2026: UPF-HNQ: ultra-processed food high nutritional quality and UPF-LNQ: ultra-processed food low nutritional quality). Positive SMD (standardised mean difference) indicates UPF trial arm has a higher score on the outcome (rated palatability of diet) than other arm, negative SMD indicates the opposite.

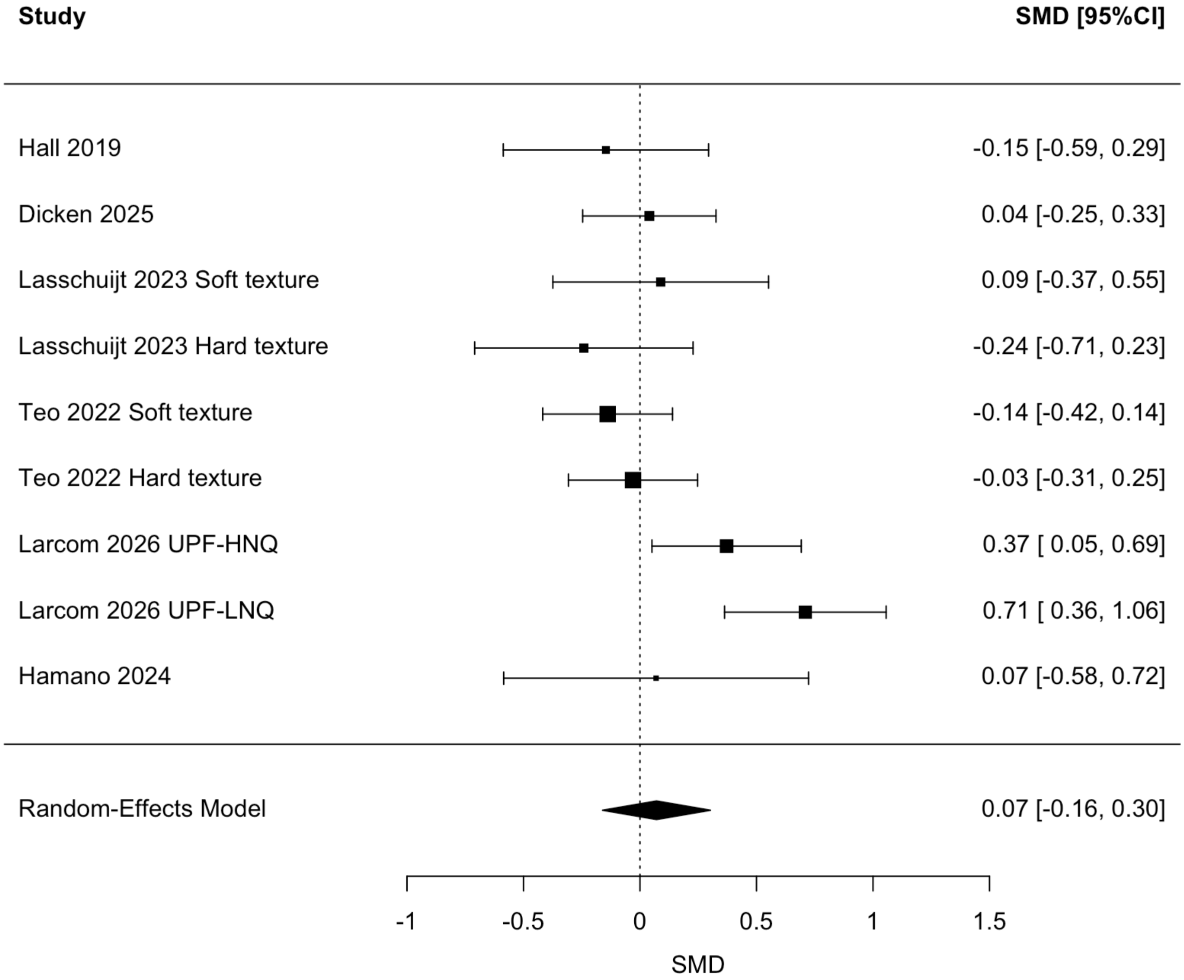


**Figure S5.** Hunger meta-analysis (Larcom 2026: UPF-HNQ: ultra-processed food high nutritional quality and UPF-LNQ: ultra-processed food low nutritional quality). Positive SMD (standardised mean difference) indicates UPF trial arm has a higher score on the outcome (hunger) than other arm, negative SMD indicates the opposite.

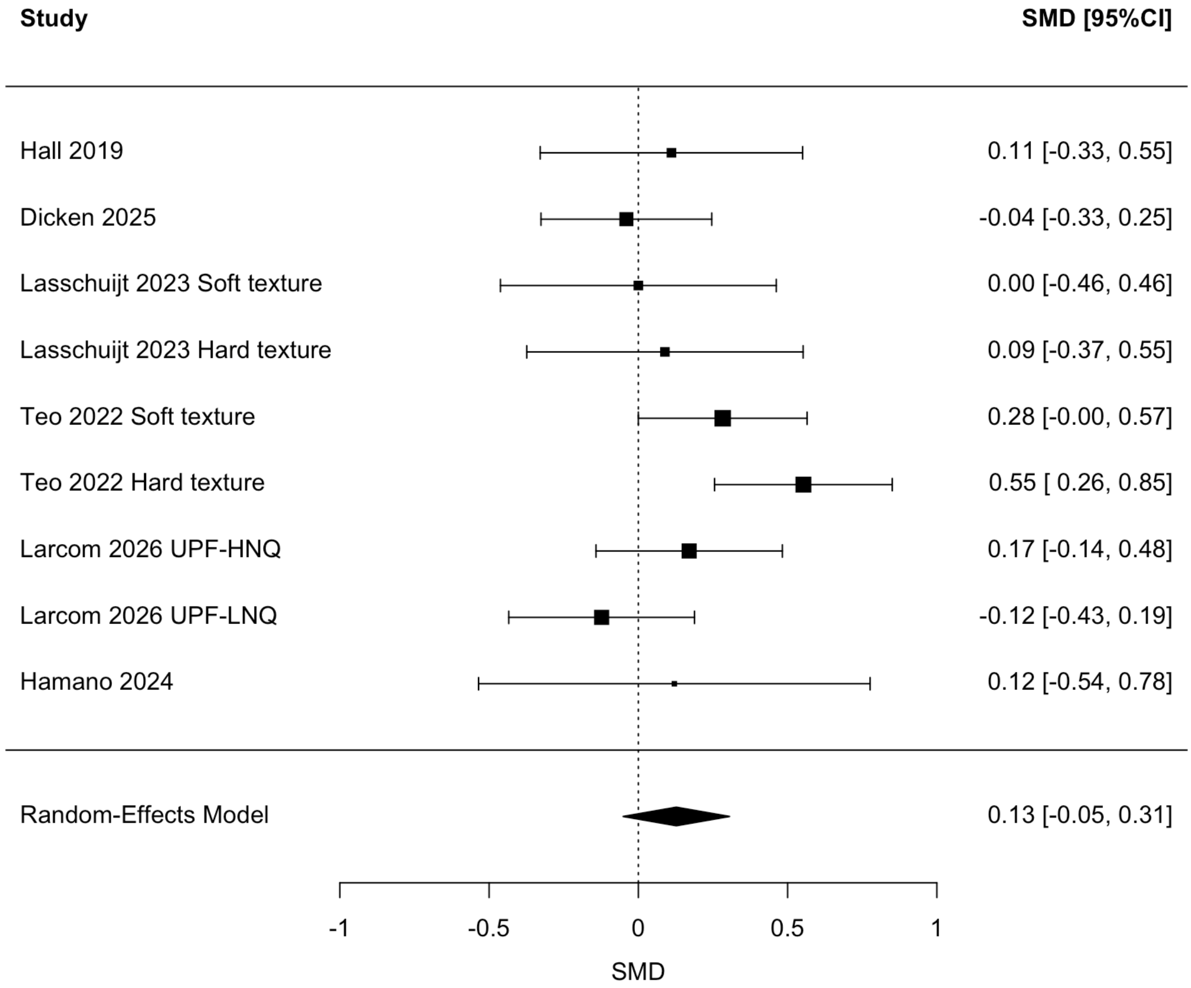


**Figure S6.** Fullness meta-analysis (Larcom 2026: UPF-HNQ: ultra-processed food high nutritional quality and UPF-LNQ: ultra-processed food low nutritional quality). Positive SMD (standardised mean difference) indicates UPF trial arm has a higher score on the outcome (hunger) than other arm, negative SMD indicates the opposite.
